# Characterizing Response to Personalized Intervention in Lower Limb Prosthesis Users: A Sensor-Based Approach to Rehabilitation

**DOI:** 10.1101/2025.07.21.25331936

**Authors:** Sara Nataletti, Jennifer Bartloff, Juan Cave, Henry Wyneken, Amber Wacek, Matthew McGuire, Shenan Hoppe-Ludwig, Anushua Banerjee, Rachel Maronati, Brad D. Hendershot, John M. Looft, Arun Jayaraman

## Abstract

**Question:** Can a personalized post-rehabilitation intervention, guided by wearable sensor data, clinical outcomes, and expert review, improve functional mobility in individuals with lower limb amputation (LLA) who exhibit unmet mobility needs?

**Design:** Prospective observational study with a nested intervention.

**Participants:** Fifty-eight adults with LLA, recruited from military and civilian sites. Based on three months of community-based monitoring and clinical assessments, an expert panel identified 26 individuals as requiring additional support; 19 completed the intervention.

**Intervention:** Tailored combinations of physical therapy, prosthetic adjustments, and/or motivational interviewing delivered over three months based on individual needs and goals.

**Outcome Measures:** Performance-based tests, patient-reported outcomes, daily step count, and progress toward self-defined mobility goals. Change from baseline was assessed using minimal clinically important difference (MCID) thresholds. Between-group comparisons (Responders vs Non-Responders), and equivalence testing with a non-intervention group were conducted.

**Results:** Following intervention, 68% of participants improved in functional outcomes, goal attainment, or both. Among Responders, 46% exceeded the MCID for sit-to-stand performance, with 17– 33% meeting thresholds for other mobility and quality-of-life measures. Responders had lower body mass index, fewer comorbidities, showed greater gains in step count, physical performance and prosthesis-related quality of life. Equivalence testing showed Amputee Mobility Predictor scores among Sustained Responders became statistically comparable to those who never required intervention. Most participants maintained gains at follow-up.

**Conclusion:** A personalized intervention approach guided by integrated data and experts review may improve real-world outcomes in individuals with LLA. These findings support reassessment to identify residual needs and deliver targeted support beyond standard care.

## Introduction

Lower limb amputation (LLA) profoundly impacts an individual’s physical conditions, significantly affecting mobility, independence, and quality of life. In the United States, over 2.3 million individuals live with limb loss, with approximately 560,000 new amputations annually, over 90% of which involve the lower extremity^1–3^. Regardless of etiology—trauma, vascular disease, and congenital conditions—LLA introduces lasting physical, psychological, and social challenges^4–7^. Rehabilitation programs aim to restore independence and support community reintegration through improved mobility, meaningful activities, and social participation^8^. However, many individuals do not achieve or sustain their full potential for community mobility despite gains made during traditional rehabilitation and access to advanced prosthetic technologies^9^.

Clinical practice guidelines (CPGs) from the Department of Veterans Affairs (VA) and the Department of Defense (DOD) advocate for structured post-rehabilitation care^10^ but their implementation in routine clinical settings, particularly in civilian systems, remains inconsistent. Limited insurance coverage, lack of follow-up protocols, and fragmented care coordination contribute to delayed identification of functional decline and missed opportunities for timely intervention^11,12^.

Currently, clinical decision-making during rehabilitation often relies on a narrow set of standardized performance tests and the Medicare Functional Classification Level (K-level) system^13^. While these ratings help guide prosthetic prescriptions and functional classifications, they are inherently limited, offering only brief and episodic snapshots of functional ability under controlled conditions, potentially constraining access to advanced prosthetic technologies and support services over time^14^. Standardized performance tests frequently overlook evolving real-world challenges, such as prosthetic discomfort, motivational barriers, environmental obstacles or patient-defined goals, factors that strongly influence long-term mobility and participation^15–17^. After discharge from rehabilitation care, individuals may gradually encounter emerging mobility limitations or disengagement, leading to reduced community integration and a mismatch between clinically expected function and daily reality.

Wearable sensors offer a valuable window into real-world activity and prosthesis use beyond the clinic^18–21^. Wearable technology studies consistently show mobility of individuals with LLA fall below recommended daily activity levels^22,23^ and demonstrate wide variability in prosthesis wear time^24,25^, revealing gaps that conventional follow-up methodologies (e.g., in-clinic assessments, self-reported questionnaires, and clinician check-ins) frequently overlook. However, to be clinically meaningful, sensor data must be interpreted in the context of structured clinical assessments, patient-reported outcomes, and individualized mobility goals. Together, these findings highlight the need for more responsive systems to detect and address mobility challenges after traditional rehabilitation ends.

To bridge these gaps, this study aimed to develop and employ a toolbox of performance-based, patient-reported, and sensor-based outcomes to: (1) comprehensively characterize real-world mobility and progress toward personal goals, (2) evaluate responses to an individually tailored intervention model, and (3) explore factors associated with improvement (or lack thereof) in comparison to those who consistently met expectations and did not require additional support. A multidisciplinary expert panel reviewed integrated data to identify individuals with unmet mobility needs and guided the delivery of personalized interventions. We hypothesized that individuals identified as having unmet mobility needs would demonstrate meaningful improvements following intervention toward functional levels similar to those who were already meeting mobility expectations. These efforts support the development of a scalable, patient-centered model to generate a multidimensional view of real-world function, thereby extending rehabilitation beyond the clinic and addressing persistent gaps in current care delivery.

## Methods

### Participants

Participants were recruited from three medical research facilities: the Shirley Ryan Ability Lab (SRALab) (Chicago, IL, Population: Civilian), Minneapolis Veterans Affairs Health Care System (MVAHCS) (Minneapolis, MN, Population: Veteran), and Walter Reed National Military Medical Center (WRNMMC) (Bethesda, MD, Population: Service Member). Participants were recruited in 2020-2022 primarily using research registries and prosthetic clinics (ClinicalTrial.gov NCT03930199). The study was approved by the Institutional Review Board of each site (SRALab: STU00208186, MVAHCS: VAM-18-00362, and WRNMMC: WRNMMC-2019-0226), and all participants provided written informed consent.

Eligible participants were individuals between 18 and 89 years old with either unilateral or bilateral LLA at the transtibial or transfemoral level, including those with knee disarticulations. They were required to have been using at least one definitive prosthesis (i.e., a stable, non-temporary prosthesis with a finalized socket and componentry unlikely to change) for a minimum of six months. Participants were required to have a designated K-level of K2 or higher, indicating the ability to ambulate within the household and manage low-level environmental barriers using a prosthesis. Exclusion criteria included co-morbidities that would limit prosthesis use, such as severe traumatic brain injury or cerebral vascular accident, as well as reluctance to engage with the study’s smartphone application.

### Study Design

Participants could complete up to four in-clinic testing visits (i.e., T-1, T0, T1, and T2), each occurring at three-month intervals. Additional visits for sensor exchange or technical support were conducted as needed (Figure 1).

**Figure 1.**
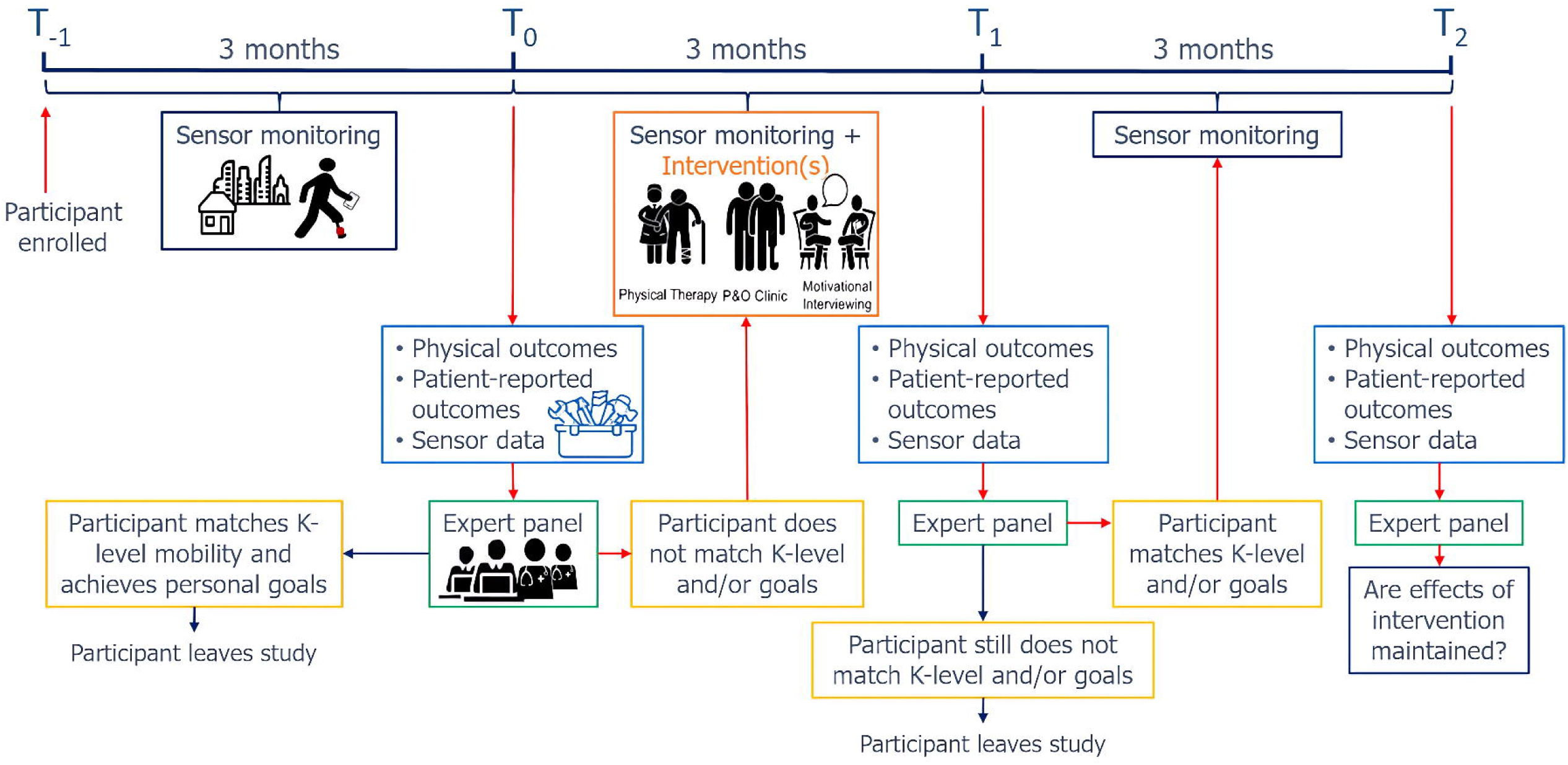
Study protocol and participant flow. After an initial three-month community mobility monitoring phase (T-1 to T0), participants completed clinical and self-reported assessments. A cross-site expert panel reviewed combined data to determine whether participants met expected mobility (based on prescribed K-level) and personal goals. Participants meeting expectations exited the study, while others received a three-month personalized intervention. At T1, a second review determined improvement status. Responders entered a final follow-up phase; Non-Responders exited the study. A third review at T2 assessed whether improvements were maintained

At enrollment (T-1), medical history and demographic information were collected, and participants were fitted with an ActiGraph wGT3X-BT (ActiGraph; Pensacola, FL, USA) tri-axial accelerometer, placed on their primary prosthesis, for continuous community mobility monitoring.

Each subsequent visit (T0, T1 and T2), included a standardized battery of clinical outcome measures, including both performance-based assessments and patient-reported outcomes. At T0, participants also completed a semi-structured interview to assess personal mobility goals, barriers to prosthesis use, and contextual factors influencing community participation. The interview combined open-ended questions with structured prompts to ensure consistency across sites while capturing individual experiences, perceived challenges and their capacity to engage in rehabilitation strategies.

After each assessment visit, community-based sensor data from the prior three-month period were summarized and integrated with performance-based and patient-reported data for review by a cross-site expert panel, in accordance with recommendations from the Centers for Medicare & Medicaid Services (CMS)^26^. The expert panel consisted of physical therapists, prosthetists, health providers and researchers from all three participating sites, with at least one representative from each discipline and each site involved in the review process^27–29^. All participant data were de-identified prior to panel review to minimize bias and ensure objective decision-making. Panel decisions were made by majority vote and served as the reference standard for intervention assignment and response classification.

At T0, the panel assessed whether the participants were achieving community activity levels consistent with their prescribed K-level and meeting their personal mobility goals. Those meeting expectations exited the study. Participants with unmet needs entered a three-month personalized intervention phase (T0–T1), receiving individually tailored treatment (e.g., prosthetic care, physical therapy, motivational interviewing) while continuing mobility monitoring with an ActiGraph.

At T1, the panel re-evaluated participants to determine improvement in clinical outcomes and/or goal attainment, categorizing them as Responders or Non-Responders. Only Responders proceeded to a final three-month, non-intervention monitoring period; Non-Responders exited the study.

At T2, the panel determined whether improvements were sustained. Participants were classified as Sustained Responders if gain were maintained, or Non-Sustained Responders if a decline or loss of progress was observed.

### Collected Outcomes

Participant demographic and medical history information was obtained through electronic health records, clinical evaluations (therapy and prosthetic) and self-reported questionnaires. Collected variables included age, sex, height, weight, body mass index (BMI), etiology of limb loss, time since amputation, level and laterality of amputation (e.g., transtibial, transfemoral, unilateral or bilateral), prescribed K-level, and presence of comorbidities.

Performance-based assessments were conducted in research or clinical settings and included the Ten-Meter Walk Test self-selected (10MWT-SS) and fast velocity (10MWT-FV), Six-Minute Walk Test (6MWT), Five-Time Sit to Stand Test (5xSTS), Four Square Step Test (FSST), Berg Balance Scale (BBS), Functional Gait Assessment (FGA), Amputee Mobility Predictor (AMP), and the Comprehensive High-Level Activity Mobility Predictor (CHAMP).

Patient-reported outcome measures included the Orthotics and Prosthetics Users’ Survey (OPUS): *Health Related Quality of Life Index* (HQOL)*, Lower Extremity Functional Status* (LEFS)*, and Satisfaction with Device and Services* (SAT); Modified Falls Efficacy Scale (mFES); Prosthesis Evaluation Questionnaire (PEQ): *Ambulation (AM), Appearance (AP), Frustration (FR), Perceived Response (PR), Residual Limb Health (RL), Social Burden (SB), Sounds (SO), Utility (UT), and Well-Being (WB), and Mobility scale combining ambulation and transfer (MS)*; Prosthetic Limb Users Survey of Mobility (PLUS-M); Patient-Reported Outcomes Measurement Information System (PROMIS): *Mobility, Physical Function with Mobility, and Depression*; Patient Health Questionnaire-9 (PHQ-9); Community Participation Indicators (CPI).

Daily step counts were calculated using ActiGraph’s proprietary regression-based (REG) step detection algorithm^30^.

### Intervention

Participants were assigned to one or more interventions—Motivational Interviewing (MI), Prosthetic Care (Px), or Physical Therapy (PT)—based on expert panel recommendations and clinical judgment at each site, informed by the T0 interview and participant mobility goals. Interventions were tailored to individual needs and delivered over a three-month period.

Participants with physical limitations, such as poor prosthesis fit, discomfort, or deconditioning, received Px and/or PT. Px included socket adjustments, component alignment, repairs, or replacements, as recommended by the clinician and within reimbursement guidelines. PT focused on gait retraining, balance, strengthening, pain management, and activities of daily living, aligned with participant goals. Frequency and duration were determined by the clinical team.

Participants whose prosthesis use, and community mobility were limited by psychological factors or barriers were referred for MI. MI was delivered by trained clinicians who completed a multi-day training program prior to implementation. The intervention included an initial 30–45 minute in-person session to build rapport and identify participant-defined goals, followed by eight structured sessions over three months, weekly in the first month, biweekly in the second, and one final session in the third. Sessions followed a standardized framework, with flexibility for clinicians to apply MI techniques to explore participants’ perspectives more deeply.

### Data and Statistical Analysis

We conducted a series of analyses to characterize the response to personalized intervention across clinical, performance-based, patient-reported, and real-world mobility outcomes. Descriptive statistics summarized the demographic characteristics of participants who received interventions, including relevant medical history and type/s of intervention they received.

To characterize overall response to interventions, we quantified the number of participants who improved in clinical outcomes, achieved or made progress toward their personal mobility goals, and/or demonstrated advancement in functional classification (i.e., prescribed K-level). Among Responders, sustained improvements were further assessed at three-month follow-up (T2). Individual changes across outcomes were visualized with circular heatmaps to illustrate response patterns. To support interpretation, mean and individual changes were compared to published minimal clinically important differences (MCIDs), and the proportion of participants exceeding MCIDs was calculated for each group.

To examine intervention response patterns, we first compared absolute outcome values between groups (Responders vs. Non-Responders) at baseline (T0) and post-intervention (T1). Two-sided (ts) Wilcoxon rank-sum tests were used for T0 comparisons to assess pre-intervention differences; one-sided (os) tests were used at T1 to evaluate expected post-intervention improvements, based on the directional hypothesis that Responders would demonstrate greater improvements following intervention. We then calculated the percent change in each outcome from T0 to T1 for each participant using the formula (T1 – T0) / T0, separately within each group. Percent change was used to provide a standardized measure of improvement relative to each participant’s baseline, allowing for more meaningful comparisons across individuals and outcomes. Between-group comparisons of percent change were conducted using one-sided Wilcoxon rank-sum tests, aligned with the hypothesized direction of change. Stability of performance from T1 to T2 was tested for Responders for each outcome using a two-sided Wilcoxon signed-rank. Categorical variables (e.g., comorbidities such as diabetes or arthritis) were compared between groups using Fisher’s Exact Test. All analyses used non-parametric tests due to the small sample size. P-values were not adjusted for multiple comparisons, as the analyses were exploratory and intended to identify patterns that may inform future studies, accepting a higher risk of Type I error.

To evaluate agreement between expert panel decisions and clinical outcome measures, change scores were calculated for each participant by comparing T1 values to baseline (T0). Each outcome was dichotomized as “Improved” (>0% change from T0) or “Not Improved” (≤0% change) and compared to expert classification (Responder vs. Non-Responder) using percent agreement and Cohen’s Kappa (κ). Statistical significance was evaluated with one-tailed z-tests (α = 0.05).

Finally, we conducted equivalence testing using a bootstrapped variation of the t-test to determine whether individuals who improved following intervention and maintained gains at T2 were functionally comparable to those who never required intervention. Analyses were restricted to participants classified as K3 by the expert panel to control for baseline mobility level. To contextualize post-intervention comparisons, equivalence test was also applied at T0 to assess their level of equivalence before the intervention and help determine whether any observed equivalence at T2 reflected meaningful change rather than pre-existing similarities or statistical artifacts. The Two One-Sided Test (TOST) procedure^31,32^ was applied using established MCID thresholds for AMP, 10MWT, 5xSTS, PLUS-M, OPUS HQOL, mFES, and PROMIS Depression. Equivalence was concluded when the 90% confidence interval for between-group differences fell entirely within the MCID-defined bounds. If the confidence interval extended beyond the equivalence margins, we interpreted this as equivalence not being established.

All analyses were conducted using R (v2024.12.1) and Python (scikit-learn, SciPy libraries). No corrections were made for multiple comparisons given the exploratory nature of the study.

## Results

Of the 66 participants enrolled, 58 completed the initial three-month monitoring phase. Twenty-six met criteria for intervention due to suboptimal mobility performance and/or unmet personal goals. Nineteen completed the intervention phase, of whom 13 showed improvement in clinical outcomes, goal attainment, or both, and advanced to the post-intervention follow-up. Six did not improve and exited the study. Of the 13 who progressed, 11 completed the final three-month monitoring period. Figure 2 presents the CONSORT flow diagram. Demographic characteristics of the 19 participants who underwent intervention are reported in Table 1.

**Figure 2.**
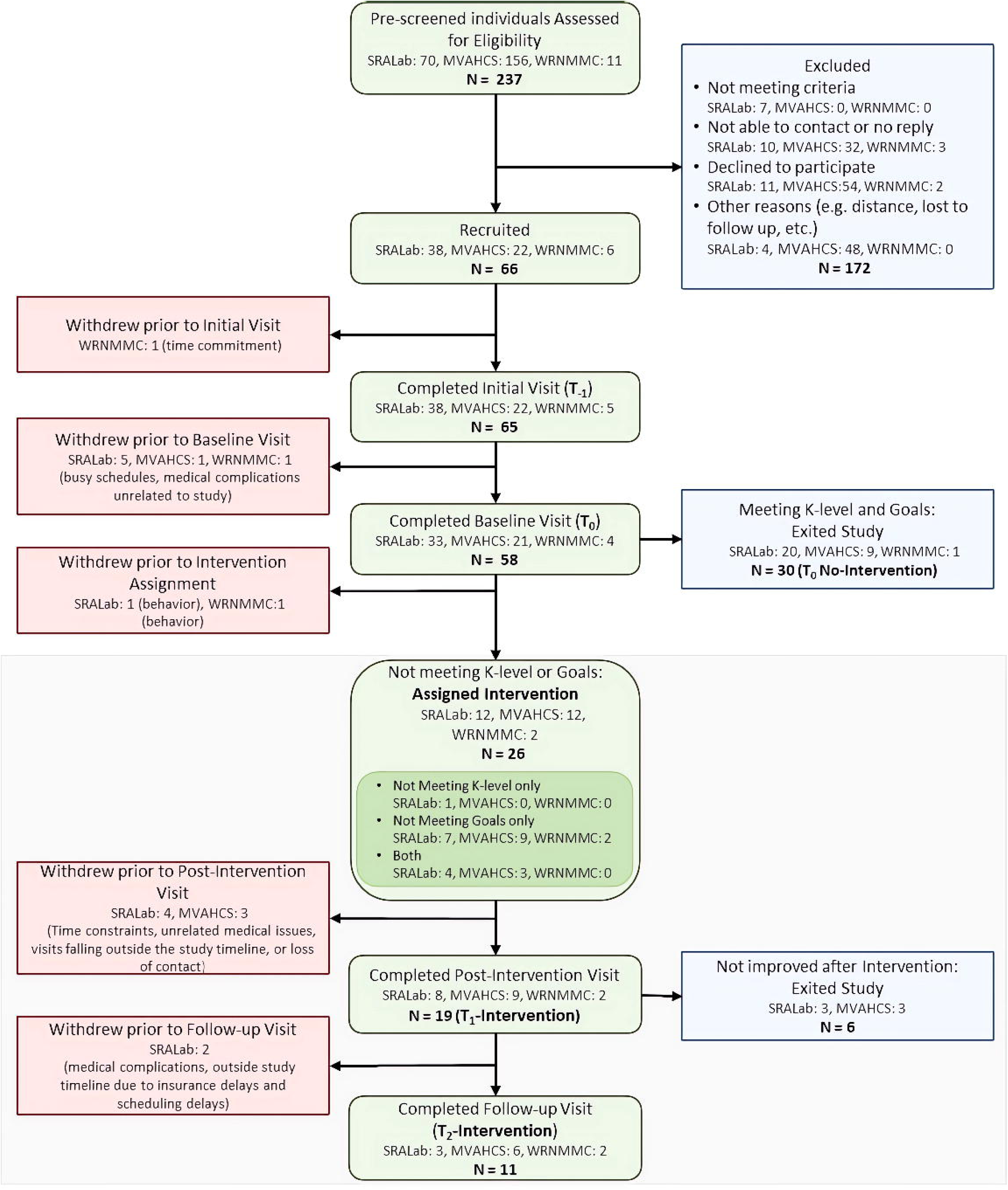
Flowchart describing the number and reason for exclusion of participants from the final data set. Abbreviations: N, number of participants; SRALab, Shirley Ryan AbilityLab; MVAHCS, Minneapolis VA Health Care System; WRNMMC, Walter Reed National Military Medical Center

**Table 1.**
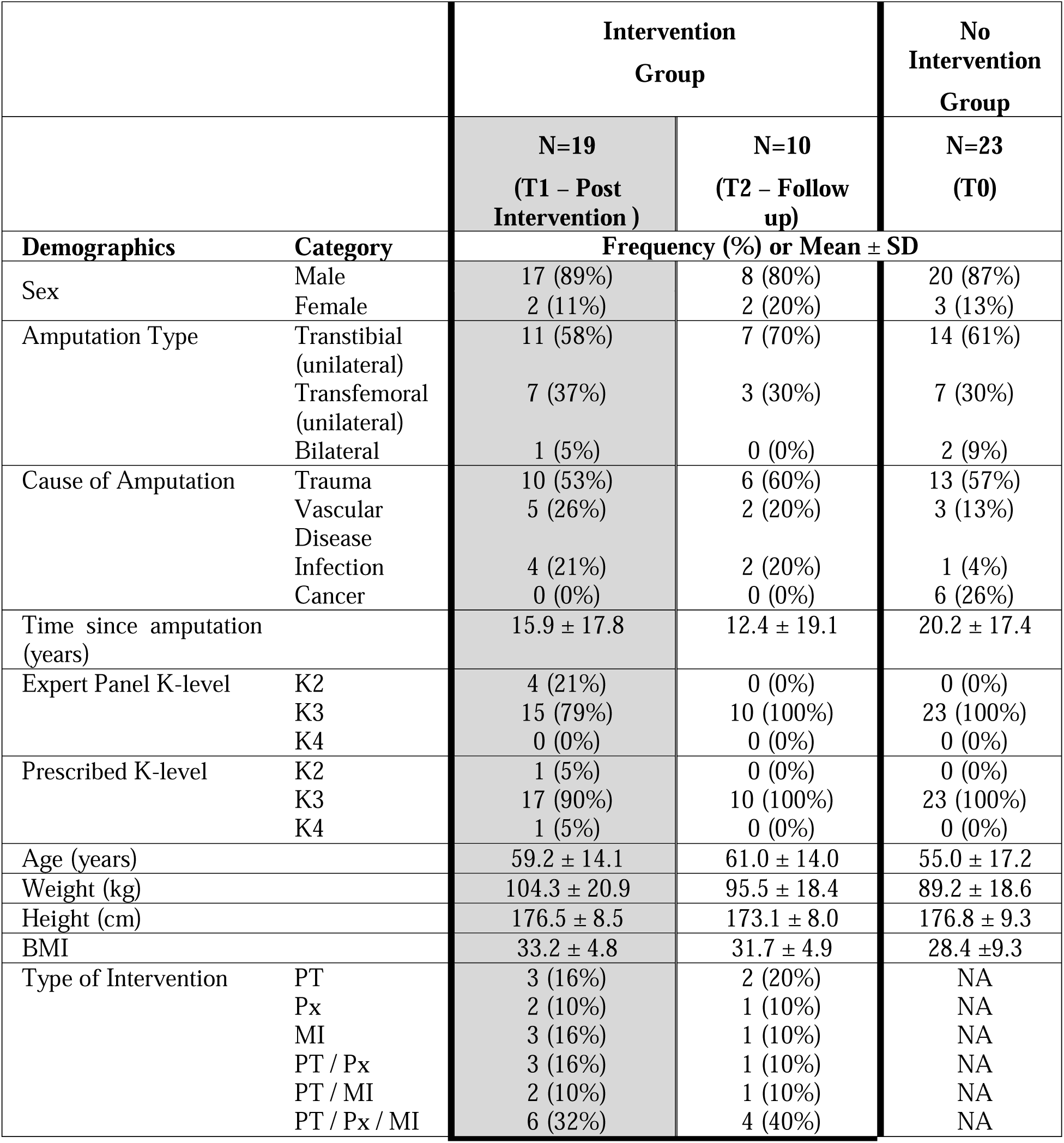
Participant demographics across study groups. Demographic characteristics of participants who completed the intervention phase (N=19), participants who achieved sustained response at follow-up (T2, N=10), and participants who did not require intervention (N=23). Demographics for T2-Intervention and T0 Non-Intervention groups are presented to support equivalence analyses. For comparability in equivalence analyses, only participants with an estimated K3 level were included, excluding those classified as K2 or K4. Among the initial 30 non-intervention participants, 23 met this criterion as did 10 of the 11 participants assessed at T2. NA = not applicable.

Following the intervention phase (T1), 13 out of 19 participants (68%) demonstrated improvements in mobility, either through enhanced clinical outcome measures, goal attainment, or both. Improvements varied, including changes in K-level and full or partial goal achievement. The remaining 6 participants (32%) were classified by the expert panel as having shown no progress in either domain. A detailed breakdown of participant flow and types of progress achieved is presented in Figure 3. Examples of individualized mobility goals and corresponding progress ratings are provided in Supplementary Table 1.

**Figure 3.**
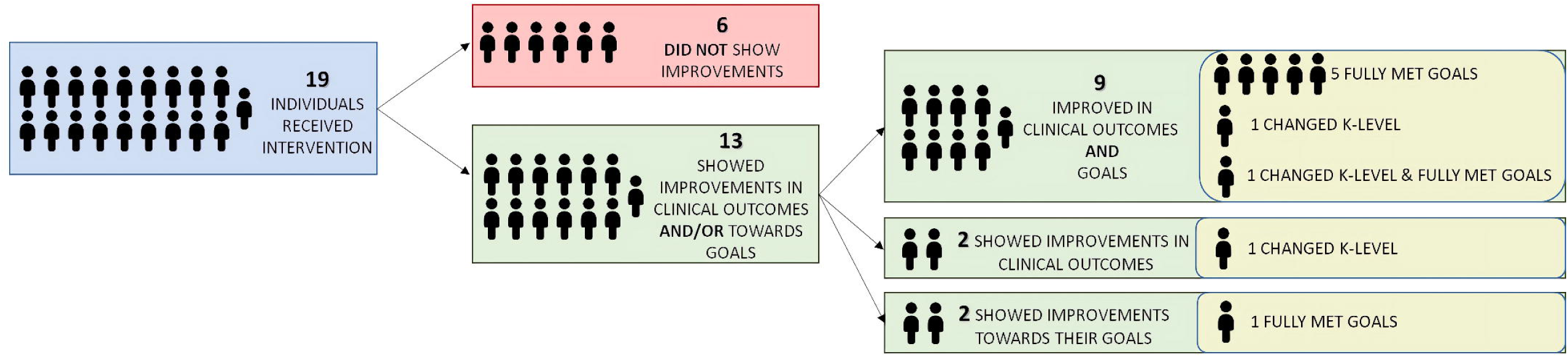
Breakdown of participant outcomes following the intervention phase (T1). Out of 19 individuals who received the intervention, 13 (68%) showed improvements in clinical outcomes, progress toward personal mobility goals, or both. Of these, 9 participants improved in both domains, while 2 improved only in clinical outcomes and 2 improved only in goal attainment. Among those who improved, some also experienced a change in K-level classification or fully met their predefined goals. Six participants (32%) did not demonstrate any measurable improvement.

Figure 4 presents circular heatmaps illustrating relative changes in performance-based, patient-reported, and mean daily step count from baseline (T0) to post-intervention (T1). Participants are grouped by expert panel classification, with Responders shown on the left and Non-Responders on the right. Each ring represents a participant, with outcome domains organized along the circumference. Colors indicate direction of change relative to baseline: green reflects improvement, gray indicates no change, and red denotes decline. These visualizations highlight the multidimensional nature of intervention effects and the variability in individual response profiles.

**Figure 4.**
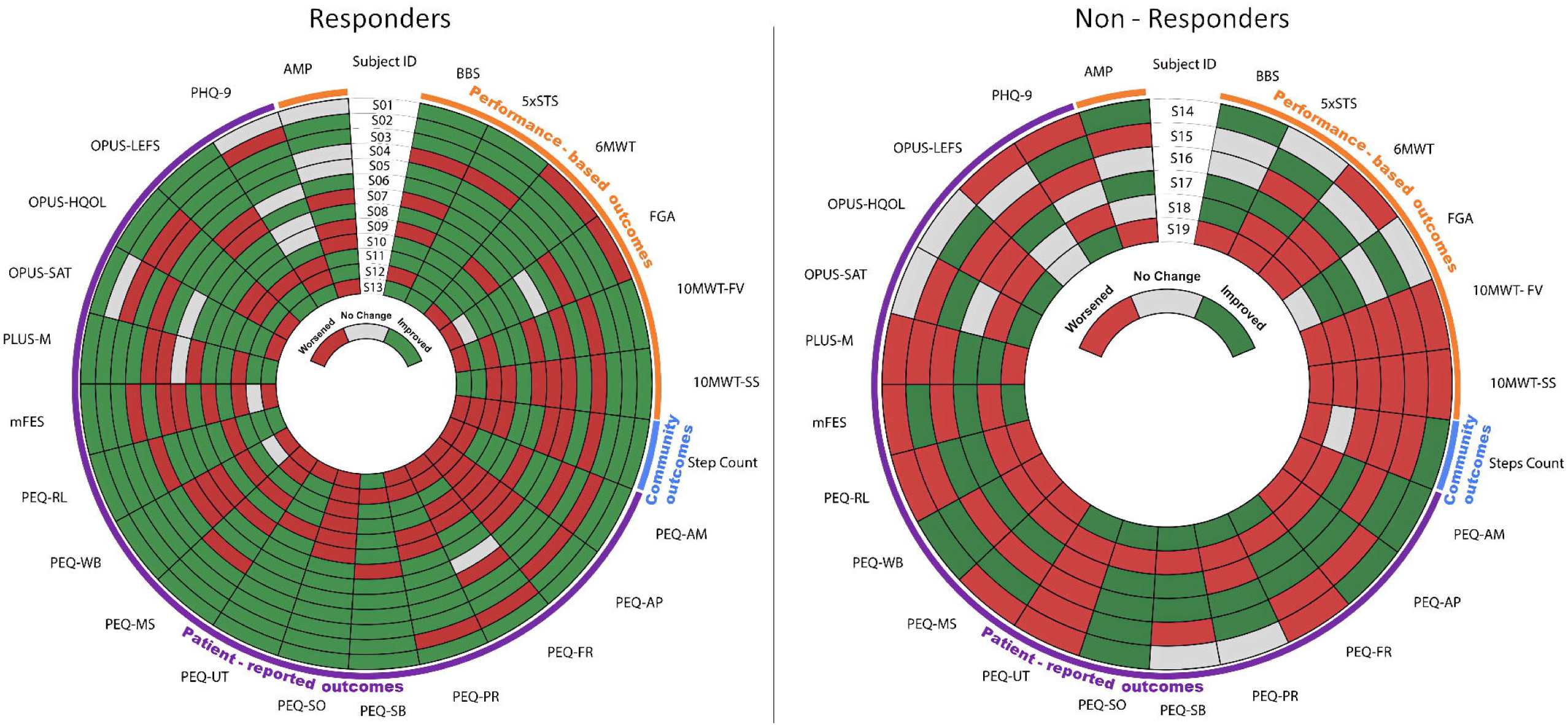
Circular heat maps illustrating changes in performance-based, patient-reported, and community-based outcomes for participants categorized by expert panel evaluation. The left panel displays participants classified as “Responders”, while the right panel shows those classified as “Non-Responders”. Each concentric ring represents an individual participant, with subject IDs shown in the innermost section. Outcome measures are arranged along the circumference. Color coding indicates post-intervention status for each outcome: green represents improvement, red indicates declined, and gray denotes no change.

While mean changes in most outcomes did not exceed published MCID thresholds, the 5xSTS demonstrated group-level improvement surpassing the MCID in the Responder group (Δ = –2.58 sec; MCID = 2.3 sec). Additionally, 46% of Responders exceeded the MCID for 5xSTS, and 17– 33% surpassed MCIDs for other outcomes, including AMP, OPUS HQOL, and 10MWT. In contrast, fewer participants in the Non-Responder group achieved clinically meaningful gains. Full group-level comparisons and individual MCID exceedance rates are presented in Table 2.

**Table 2.** MCID-Based Analysis of Clinical Outcomes. Mean change scores and percentage of participants exceeding the minimal clinically important difference (MCID) for each outcome measure, stratified by responder status. Positive or negative mean values reflect the direction of improvement for each specific measure. The percentage of participants exceeding the MCID reflects the proportion of individuals in each group (Responder and Non-Responder) who demonstrated clinically meaningful change.

| Outcome Measure | Mean $\Delta$ (Responder) | Mean $\Delta$ (Non-Responder) | MCID | % Responder >MCID | % Non-Responder >MCID |
| --- | --- | --- | --- | --- | --- |
| 10MWT-SS | 0.02 | -0.07 | 0.13 m/s<sup>32</sup> | 30% | 0% |
| AMP | 1.00 | 1.83 | 3.40<sup>33</sup> | 15% | 17% |
| 5xSTS | -2.58 | 1.67 | 2.30 s<sup>34</sup> | 46% | 17% |
| PLUS-M | 0.29 | -0.88 | 6.42<sup>35</sup> | 0% | 0% |
| OPUS-HQOL | 2.44 | -0.50 | 9.20<sup>33</sup> | 15% | 0% |
| mFES | -0.14 | -0.13 | 1.50<sup>36</sup> | 15% | 17% |
| PROMIS Depression | -1.10 | 2.25 | 6.71<sup>35</sup> | 8% | 17% |

### Demographic and Health history

Health history comparisons between Responders and Non-Responders were conducted for all participants who completed the T1 assessment, excluding one Non-Responder with unavailable health history data. Figure 5 summarizes the prevalence of reported medical conditions by group. Diabetes was significantly more common among Non-Responders (80%) compared to Responders (23%) (p = 0.047). Similarly, non-rheumatoid arthritis was reported in 80% of Nonresponders and 8% of Responders (p = 0.008). No significant differences were found between Responder and Non-Responders for other conditions, including hypertension, depression/anxiety, pain, circulation problems, fractures, and cancer (all p > 0.05).

**Figure 5.**
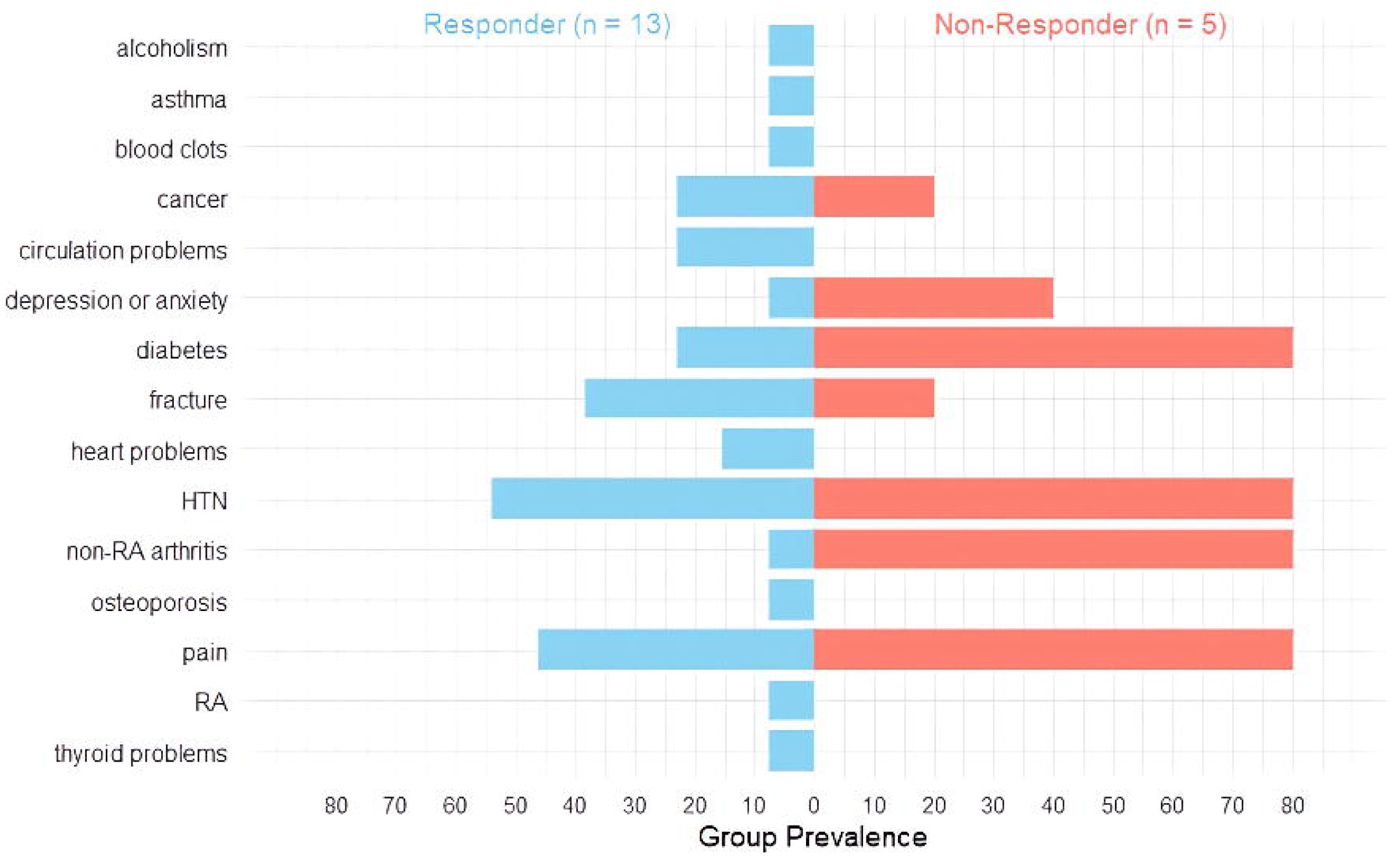
Health history among Responders and Non-Responders. Group prevalence of various conditions is shown.

BMI at T0 was significantly higher among Non-Responders compared to Responders (p = 0.024). Non-Responders were predominantly classified in the obese range (≥ 30 kg/m²), while Responders spanned a broader range, including overweight (25.0–29.9 kg/m²) or in the lower-obese categories (30.0–34.9 kg/m²) (Fig. 6).

**Figure 6.**
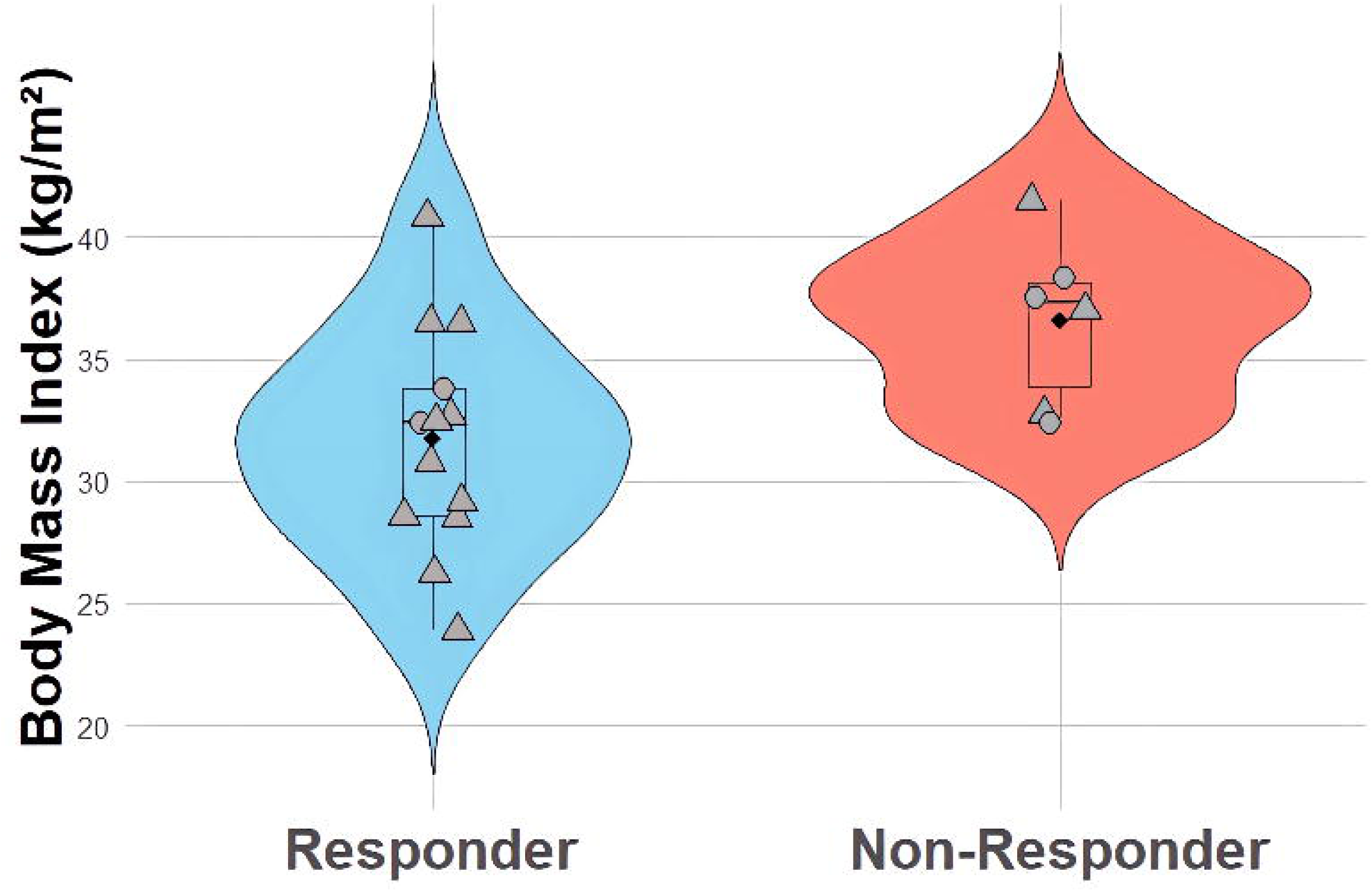
Body Mass Index (BMI) distributions at T0 by response group. Violin plots show density, with embedded boxplots and back diamonds indicating group means. Grey dots indicate individual participant values

Among participants with traumatic amputations, 8 out of 10 were classified as Responders (80%), compared to 56% (5 of 9) among those with non-traumatic etiologies (Fig. 7). This reflects a 25% absolute difference in Responder prevalence between the two groups. Individuals with traumatic amputations had higher odds of being Responders compared to those with non-traumatic etiologies (odds ratio=3.2).

**Figure 7.**
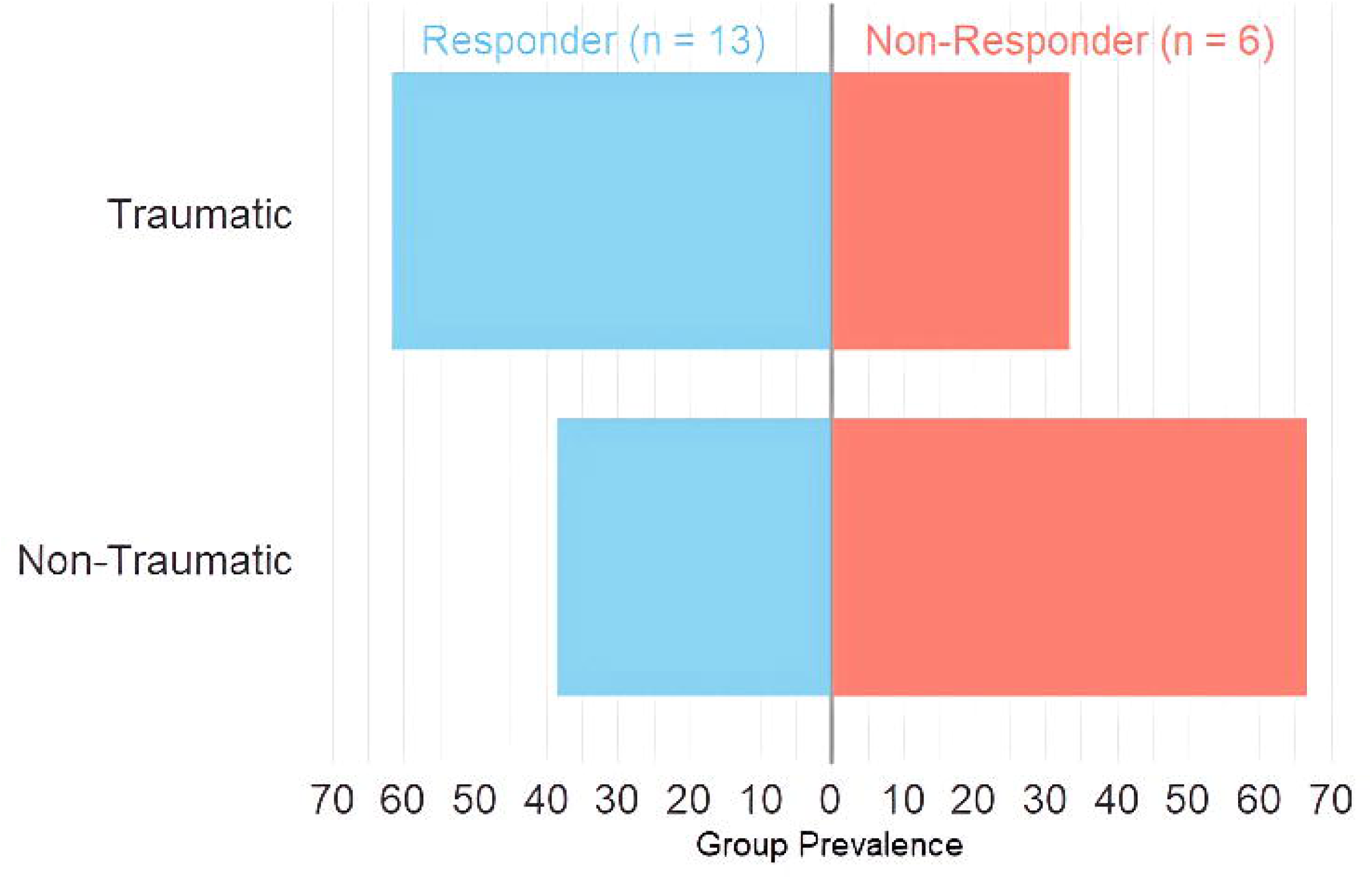
Etiology of limb loss among Responders and Non-Responders: The proportion of participants with traumatic versus non-traumatic amputation is shown for each group. Numeric labels indicate the number of participants in each category.

### Performance-Based Outcomes

No statistically significant between-group (Responder vs. Non-Responder) differences were observed on performance-based outcomes (T0; all *p* > .05, ts). At post-intervention (T1), however, Responders demonstrated significantly better performance on the 6MWT (*p* = 0.013, os), BBS (*p* = 0.021, os), 10MWT-FV (p = 0.036, os), 10MWT-SS (p = 0.029, os) and lower times on the FSST (p = 0.041, os).

When evaluating percent changes from T0 to T1, significant group differences were observed for the 6MWT (p = 0.003, os), 10MWT-FV (p = 0.006, os), 10MWT-SS (p = 0.036, os) and the FSST (p = 0.01, os). Compared to Non-Responder, Responders demonstrated greater improvements in walking speed, endurance, and dynamic balance/agility.

### Patient-Reported Outcomes

No statistically significant differences in patient-reported outcome measures were observed between Responders and Non-Responders at baseline (T0). However, at post-intervention (T1), Responders reported significantly higher scores on multiple PEQ subscales, including PEQ-MS, PEQ-AM, PEQ-FR, PEQ-SB and PEQ-WB (all p < 0.05, os).

When evaluating percent changes from T0 to T1, significant group differences were observed for the PEQ-RL (p = 0.010, os) and the PEQ-UT (p = 0.018, os).

### Step Count

To account for potential baseline differences, changes in daily step counts were normalized relative to baseline (T0) values and expressed as percent change. As shown in Figure 8, Responders exhibited a broader and more variable distribution of percent change in daily steps compared to Non-Responders, who showed a narrower and more consistently negative range of changes. Responders demonstrated greater improvements than Non-Responders in daily step count (p = 0.03, Wilcoxon rank sum exact test). Although absolute step counts did not differ significantly between groups at baseline (T0), they were significantly higher in Responders at T1 (p = 0.007, Wilcoxon rank sum exact test).

**Figure 8.**
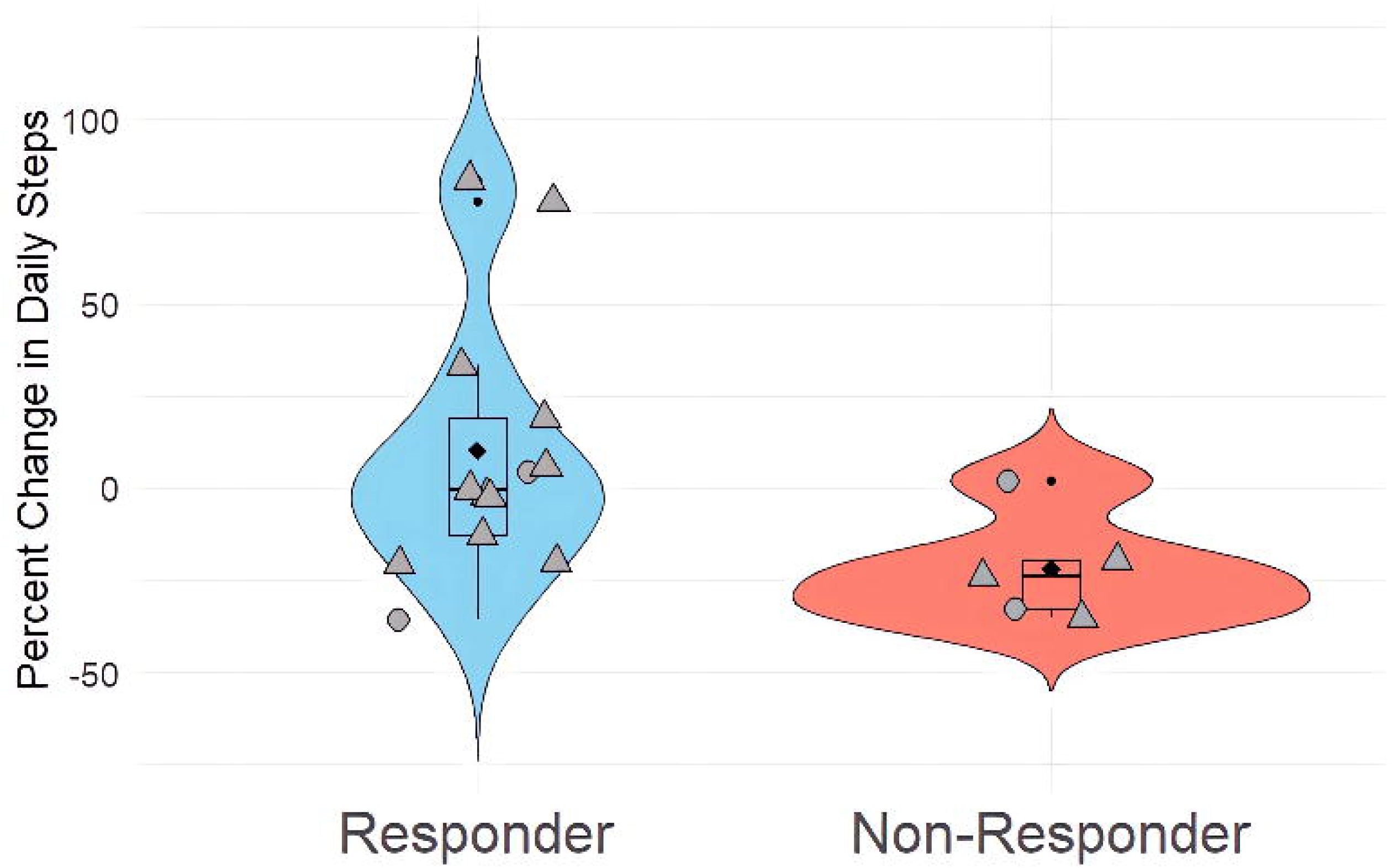
Percent change in daily steps between T0 and T1. Violin plots display the distribution of percent change in daily step counts (default filter) from baseline (T0) to post-intervention (T1) for Responders (blue) and Non-Responders (red). Each point represents an individual participant, with shapes indicating K-level at T1 (circles = K2; triangles = K3)

### Expert Panel Agreement

Agreement between expert panel classifications and observed outcome trends varied across measures. Seven out of 24 outcomes demonstrated statistically significant agreement beyond chance (p < 0.05), with Cohen’s Kappa values ranging from 0.424 to 0.650, indicating moderate to substantial agreement (Table 3). The highest agreement was observed for PEQ-UT (κ = 0.650, p = 0.0002), followed by FSST (κ = 0.557, p = 0.0084), 6MWT (κ = 0.553, p = 0.0027), and OPUS-LEFS score (κ = 0.553, p = 0.0027). Additional significant agreement was found for PEQ-RL, 10MWT-FV, and 10MWT-SS measures.

**Table 3.** Outcome Measures with Significant Agreement with Expert Panel Decisions. Percent agreement, Cohen’s kappa (κ), and p-values are reported for outcomes demonstrating significant alignment (p < 0.05) with expert panel assessments of participant improvement

| Outcome Measure | N | % Agreement | Cohen's $\kappa$ | p-value |
| --- | --- | --- | --- | --- |
| PEQ-UT | 19 | 84.21% | 0.650 | 0.0002 |
| FSST | 18 | 83.33% | 0.557 | 0.0084 |
| SMWT | 19 | 78.95% | 0.553 | 0.0027 |
| OPUS-LEFS | 19 | 78.95% | 0.553 | 0.0027 |
| PEQ-RL | 19 | 78.95% | 0.513 | 0.0089 |
| 10MWT-FV | 19 | 73.68% | 0.503 | 0.0044 |
| 10MWT-SS | 19 | 68.42% | 0.424 | 0.0146 |

### Follow-Up Evaluation (T2)

Of the 13 subjects improved after the intervention (Responders), only 11 completed the three-month follow-up period. Two participants exited the study before T2 due to unrelated medical complication and insurance delay.

At T2, 91% (10/11) of participants maintained their post-intervention improvements in clinical outcome measures. Of the four participants who had not fully met their personal mobility goals at T1, three went on to achieve full goal attainment by T2. One participant (9%) did not maintain improvement and was also the only individual who had not fully achieved their personal mobility goals, despite continuing to meet their prescribed K-level. The participant had set multiple ambitious goals (e.g., golfing, traveling, and scuba diving) and showed partial progress, however goal attainment was impacted by a pelvic fracture sustained during the follow-up phase. All participants retained or regain their prescribed K-level, with no observed decline in functional classification. Between T1 and T2, two patient-reported outcomes changed significantly: satisfaction with the prosthetic device decreased (*OPUS Satisfaction with Device Score*, *p* = 0.041, ts, effect size = –0.708), while perceived control over community participation increased (*CPI Control Score*, *p* = 0.046, ts, effect size = 0.685).

### Equivalence Testing Results (T0 vs T2)

To explore whether participants who sustained improvements following intervention reached functional levels comparable to those who never required intervention, we compared 10 Sustained Responders at T2 with 23 Non-Intervention participants at T0. All individuals included were classified as K3 by the expert panel to ensure comparable baseline functional status. Summary demographic characteristics for these two subgroups are provided in Table 1.

The analysis focused on a subset of outcome measures for which published MCID thresholds were available, including: 10MWT-SS (MCID = 0.13 m/s)^32^, AMP (3.4 points)^33^, 5xSTS (2.3 sec)^34^, PLUS-M (6.42)^35^, OPUS-HQOL Rasch (9.2)^33^, mFES (1.5)^36^, and PROMIS Depression (6.71)^35^.

At baseline (T0), equivalence could not be established between the intervention and non-intervention groups on any of the selected outcome measures, as the confidence intervals for between-group differences extended beyond the MCID bounds.

At follow-up (T2), among all tested outcomes, only the AMP score met criteria for statistical equivalence (Fig. 9). No other outcomes demonstrated statistical equivalence, and several comparisons yielded inconclusive results due to wide confidence intervals extending beyond the equivalence margins. Figure 10 presents results for both the T0 and T2 comparisons to illustrate these changes over time.

**Figure 9.**
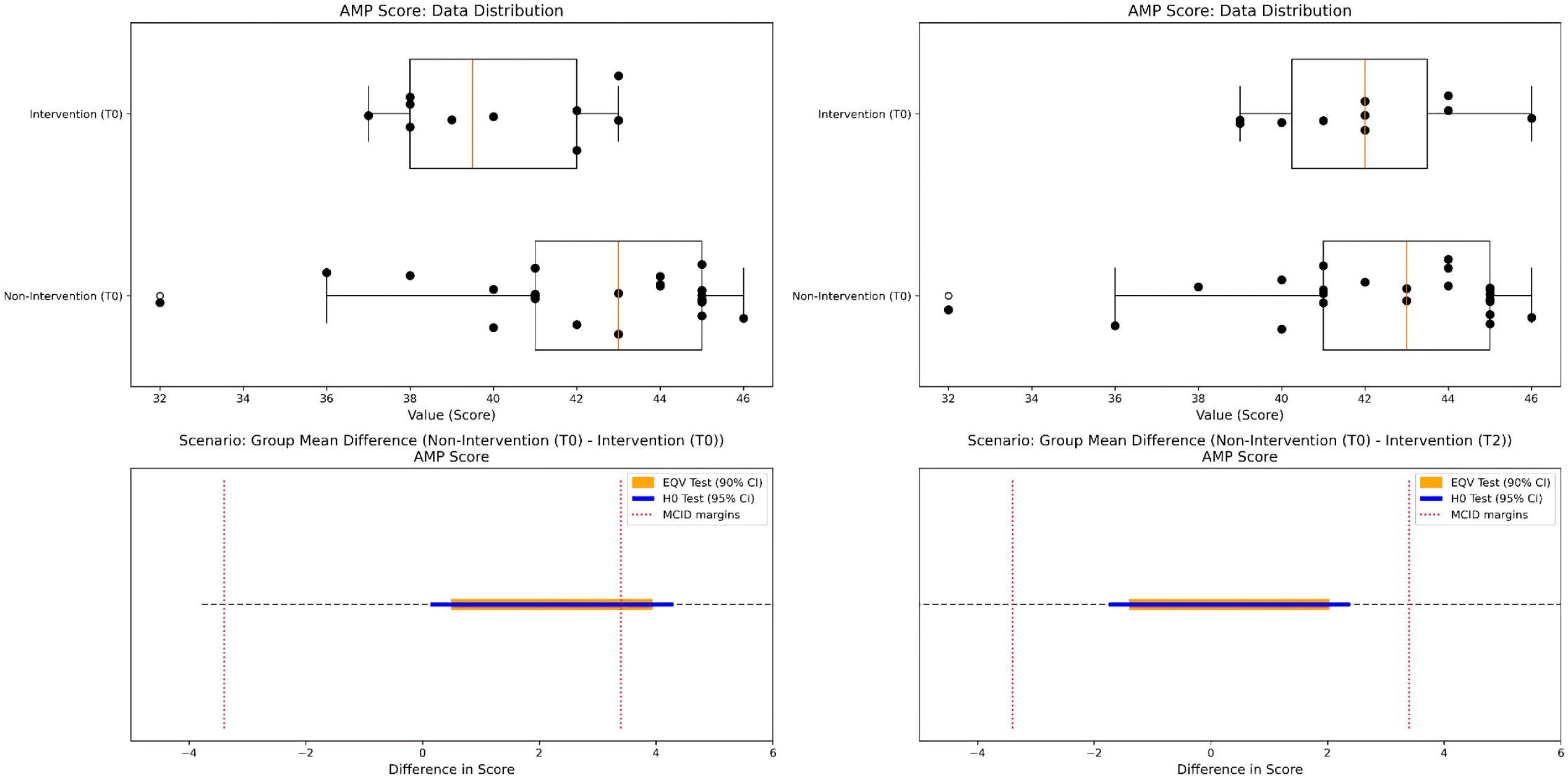
Equivalence test results for the AMP score between T2 Sustained Responders and T0 Non-Intervention participants. The top panel displays the distribution of AMP scores for each group Non-Intervention and Intervention. At baseline (T0), equivalence could not be established, as the 90% confidence interval for the between-group difference extended beyond the predefined equivalence bounds (±3.4 points). At follow-up (T2), the bottom panel shows the group mean difference (T0 – T2) and the associated 90% confidence interval (orange) used for the equivalence test, overlaid with the 95% confidence interval (blue) from a standard null hypothesis test. At T2, the 90% confidence interval fell entirely within the MCID-defined bounds, supporting the conclusion that the two groups were clinically equivalent on AMP scores.

## Discussion

This study aimed to characterize individual responses to a personalized, post-rehabilitation intervention strategy in individuals with LLA. By integrating wearable sensor data with performance-based assessments, patient-reported outcomes, and individualized mobility goals, we sought to reassess participants’ real-world function and identify those who may benefit from additional, tailored support following rehabilitation.

Among participants who received the intervention, 68% demonstrated improvements in clinical outcomes, mobility goal progress, or both. Three advanced their functional classification, and seven fully achieved previously unattainable goals. Notably, three individuals continued to make progress even after the intervention ended, ultimately attaining their goals during the follow-up phase. This suggests that the effects of personalized intervention may extend beyond the structured care period, supported by greater self-efficacy, enhanced goal awareness, and the sustained application of learned strategies. These findings align with prior research demonstrating that individualized, post-rehabilitation interventions can significantly enhance real-world mobility metrics in individuals with LLA^37,38^. Furthermore, they reinforced the value of comprehensive reassessment in identifying ongoing needs not captured by routine follow-up or standard performance testing alone^39,40^.

Outcome changes were evaluated in relation to published MCIDs. Notably, the 5xSTS demonstrated a mean improvement in the Responder group that exceeded established MCID thresholds, indicating a significant functional gain at the group level^41,42^. While group means for AMP, OPUS HQOL, and 10MWT did not consistently surpass MCIDs, a notable proportion of individuals, especially Responders, achieved clinically meaningful gains, reflecting meaningful personal progress that may be obscured in averaged results^44–46^. This pattern aligns with prior work showing that individualized, goal-oriented rehabilitation often produces heterogeneous improvements not fully reflected in group statistics^33,46^.

Importantly, baseline characteristics appeared to influence intervention response. Non-Responders were more likely to present with higher BMI, diabetes, and non-rheumatoid arthritis–conditions that have been previously associated with reduced rehabilitation outcomes and greater barriers to mobility maintenance^47^. In contrast, individuals with traumatic amputations were significantly more likely to respond positively, consistent with prior studies showing that traumatic amputees often have greater residual function and rehabilitation potential compared to those with dysvascular or other non-traumatic causes^48^. These findings suggest that baseline health and amputation characteristics may play a key role in predicting intervention response and highlight the need to tailor rehabilitation strategies to individual profiles.

At T1, Responders showed significantly greater absolute and percent-change improvements in daily step count, walking speed, endurance, and functional stability, and significantly greater absolute (but not percent-change) improvements in balance. No between-group differences were present at baseline. This supports the interpretation that these performance gains were driven by the intervention itself rather than pre-existing disparities and reinforces the validity of the expert panel’s post-intervention classification. Although not all outcome measures reached statistical significance, the consistent group differences across key mobility domains reinforce the value of expert-informed classification in identifying meaningful recovery patterns and determining who benefits most from personalized rehabilitation approaches.

Responders also showed significantly greater percent improvements in two patient-reported outcomes compared to Non-Responders: perceived ease of prosthesis use and residual limb comfort. These results suggest that functional gains following the intervention were accompanied by meaningful improvements in perceived prosthesis-related quality of life. However, by follow-up, participants reported a reduced satisfaction with the prosthetic device, whereas their perceived control over community participation had increased, highlighting the evolving nature of patient experiences after structured intervention. These shifts may reflect changing expectations or the need for continued support to sustain gains in the community, a phenomenon also observed in qualitative studies examining post-rehabilitation adjustment^50^. The reduction in device satisfaction may reflect decreased accountability and engagement once participants were no longer closely interacting with the study team. Prior studies have shown that prosthesis wear time often decreases after discharge^51^ and sedentary behavior may increase^52^, suggesting that without ongoing monitoring and reinforcement, some individuals may gradually revert to old habits or experience the reemergence of barriers identified at baseline.

The agreement analysis showed expert panel judgments aligned moderately to substantially with outcome trends for measures like PEQ-UT, FSST, 6MWT, OPUS-LEFS, PEQ-RL, and 10MWT metrics. These findings are consistent with prior work demonstrating that both performance-based measures, such as the 6MWT and FSST, and patient-reported outcomes, such as the PEQ-UT and OPUS-LEFS, can sensitively detect functional improvements following rehabilitation in individuals with LLA^54–59^. Moreover, our findings suggest that these outcome tools could help guide clinical judgments and potentially support future automated or simplified decision-making pathways.

Equivalence testing further explored whether participants who improved and maintained gains at follow-up reached comparable outcomes to those who never required intervention. At baseline (T0), no outcome measures demonstrated equivalence between these two groups. At follow-up (T2), only the AMP met statistical equivalence criteria, suggesting that some individuals initially needing support reached similar functional mobility levels to their peers after a targeted intervention. These results reinforce the clinical utility of the AMP as a sensitive tool for tracking meaningful recovery and identifying individuals approaching functional benchmarks over time^41,59^.

### Study Limitations

Several limitations must be acknowledged. While the overall study enrolled a relatively large and diverse sample, including civilian, military and veteran participants, the subgroup requiring and receiving intervention was smaller, which limited the ability to detect more subtle effects within that group. Data collection occurred during COVID-19 pandemic, which likely impacted the overall recruitment and contributed to the smaller intervention subgroup. The absence of a randomized control group limits causal conclusions about intervention effectiveness; future studies might consider reassessing non-intervention participants after a comparable follow-up period. While wearable sensor data provided valuable insights into daily activity, the analysis was limited to step count–derived features. Other aspects of community mobility, such as travel patterns captured via GPS, could not be evaluated due to missing data. The relatively long time since amputation (median 16 years) may also influence generalizability to individuals earlier in their rehabilitation journey. Additionally, outcome measures were not collected at the baseline (T-1), limiting baseline comparisons across all participants, including those who later exited the study. Finally, although the expert panel process enhanced decision-making rigor, such multidisciplinary review may not be feasible in all clinical settings. Despite these limitations, the outcome measures and clinical indicators identified in this study may inform future development of automated decision-support tools, including machine learning approaches, to help identify individuals most likely to benefit from targeted rehabilitation interventions.

## Conclusion

This study provides preliminary evidence that individualized follow-up care, guided by real-world data collected beyond traditional clinical settings, may help close the gap between functional potential and actual mobility in individuals with LLA. Reconnecting with patients after formal rehabilitation can enhance health, functional performance, and quality of life by addressing barriers that emerge during community reintegration. Periodic reassessment using ecologically valid monitoring enables clinicians to identify evolving challenges and deliver timely, targeted support. These findings support the need for scalable care models incorporating structured monitoring and personalized interventions to optimize long-term outcomes.

## Data Availability

All data produced in the present study are available upon reasonable request to the authors

## Ethics approval

This study was approved by the Institutional Review Boards of all participating sites: Shirley Ryan AbilityLab (STU00208186), Minneapolis VA Health Care System (VAM-18-00362), and Walter Reed National Military Medical Center (WRNMMC-2019-0226). All participants provided written informed consent prior to participation.

## Acknowledgements

We would like to acknowledge Jonathan R. Gladish, MS; Clare A. Severe, MS; and Stephanie Rigot, PT, PhD for their significant contributions in data collection. We would also like to acknowledge Ashley Knight, PhD and Christopher L. Dearth, PhD at WRNMMC, as well as Matthew Sauerbrey, PTA at MVAHCS for their contributions to project management. We are grateful to the members of the Expert Panel, including the clinicians and researchers that participated throughout the study at all three sites. Lastly, we would be unable to complete this work without the support of the participants willing to contribute their time and effort in collection of data for this investigation

## Author Contributions

S.N., S.H.L, M.M., A.B., J.C., A.W, conducted data collection. S.N., J.B., R.M., H.L.W. analyzed and interpreted data. S.N., S.H.L, A.B. conducted literature research. B.H., J.M.L., and A.J. were involved in the conceptualization of the project, protocol development, and funding acquisition. S.N. wrote the initial draft of the manuscript. All authors contributed to reviewing and editing the final draft. All authors read and approved the final manuscript.

