## Supplemental Table for "Characterizing Response to Personalized Intervention in Lower Limb Prosthesis Users: A Sensor-Based Approach to Rehabilitation"

**Supplementary Material**

**Examples of Individualized Mobility Goals and Progress Ratings**

This table presents a sample of individual mobility goals defined by participants at baseline (T0), along with their observed progress at follow-up. The goals reflect each participant’s personal priorities and functional aspirations, illustrating the diversity of intervention targets across the cohort. Progress toward each goal was categorized as “Fully met,” “Toward” (partial progress), or “No progress,” based on qualitative reports and clinical assessments. These examples provide context for the individualized approach used in the study and demonstrate how progress was evaluated in relation to participant-defined outcomes.

**Supplementary Table 1. Personal mobility goals defined at T0 and corresponding progress following the intervention**. Each participant's self-identified goals were documented at baseline, and progress was assessed post-intervention. Progress is categorized as **“Fully met,”** **“Toward”** (partial progress), or **“No progress.”** Green shading indicates participants who showed improvement (either fully met or partial progress), while red shading denotes participants who did not show any improvement in goal attainment. "NA" indicates that no personal goals were reported by the participant.

| **Personal Goals defined at T0** | **Progress** |
| --- | --- |
| Stair navigation, tidying apartment using prosthesis | No progress |
| NA (no personal goals) | NA |
| Run, participate in adaptive sports. | Fully met |
| Cycling, be "more nimble", descend stairs reciprocally, walk on uneven ground | No progress |
| Increase standing tolerance to be able to cook/bake and participate in more activism, take dog for walk, increase walking endurance with and without spc | No progress |
| Walking dog, going to the gym and walking on the treadmill, biking, rollerblading, traveling, skydiving/ziplining | Fully met |
| Do free weight squats instead of on smith machine | Toward |
| Reducing pain when doing physical activity | Fully met |
| Leg weight exercises, cycling, adaptive sports (hand cycling Paralympic goals), running | Toward |
| Walk and do yardwork around water on slippery and uneven terrain. Walk long distances without cane. | Fully |
| Hiking, entering/exiting kayak, returning to work part-time | No progress |
| Increasing activity levels, lose weight, floor transfers, difficulty on terrain | Toward |
| Walking longer distances, hiking, signing up for the mentor program at the VA, do home repairs and garden. | Fully |
| Limiting pain when wearing their prosthesis and walking | No progress |
| Decrease pain during prolonged prosthesis use, ability to walk long distances, and physical activity without pain or with reduced pain. | Fully |
| Walk longer distances, more natural gait, walk without an assistive device, better posture (decrease back pain) | No progress |
| Increase endurance to play golf longer, walk further, better at stairs. | Fully |
| Get stronger, more agile, more flexible, and more confident in wearing his prosthesis | No progress |
| Would like to walk further and better, spend more time in the yard, and stand longer to cook. | Toward |
